# Spleen stiffness can predict liver decompensation and survival in patients with cirrhosis

**DOI:** 10.1101/2022.10.12.22281009

**Authors:** Dimitrios S Karagiannakis, Theodoros Voulgaris, George Markakis, Dimitra Lakiotaki, Elisavet Michailidou, Evangelos Cholongitas, George Papatheodoridis

## Abstract

**Background/Aim:** Liver stiffness measurement (LSM) has been predicting liver decompensation and survival in cirrhotics. The aim of our study was to investigate if spleen stiffness measurement (SSM) by 2D-Shear Wave Elastography could predict better the probability of decompensation and mortality, compared to LSM and other parameters.

**Methods:** Consecutive cirrhotic patients were recruited between 1/2017-12/2021. LSM and SSM were performed at baseline and epidemiological, clinical and laboratory data were collected. Clinical events were recorded every 3 months.

**Results:** Totally, 177 patients were followed for a mean period of 31±18 months. In Cox regression analysis, only SSM was independently associated with the probability of decompensation (HR: 1.063, 95% CI: 1.009-1.120; p=0.021), offering an AUROC of 0.710 (p=0.003) for predicting 1-year liver decompensation (NPV: 81.1% for the cut-off point of 37 kPa). The occurrence of death/liver transplantation was independently associated only with higher SSM (HR: 1.043; 95% CI:1.003-1.084; p=0.034). The AUROC of SSM for predicting 1-year death/liver transplantation was 0.72 (p=0.006), (NPV: 95% for the cut-off of 38.8 kPa). The performance of SSM to predict the 1-year death/liver transplantation increased in high-risk patients (CTP: B/C plus MELD >10 plus LSM >20 kPa) giving an AUROC of 0.80 (p<0.001). Only 1/26 high-risk patients with SSM <38.8 kPa died during the first year of follow-up (NPV: 96.4%).

**Conclusions:** SSM was the only factor independently associated with the probability of decompensation and occurrence of death, showing better diagnostic accuracy for the prediction of 1-year decompensation or death compared to LSM and MELD score.

## Introduction

Patients with liver cirrhosis have an increased risk for poor outcome due to complications associated with liver failure and portal hypertension (PH) [1]. Hepatic venous pressure gradient (HVPG) has been considered as the reference standard method for the evaluation of PH [2], as its accuracy to predict esophageal varices formation, risk of variceal bleeding, disease decompensation and increased mortality has been well validated (3-8). Nevertheless, HVPG is an invasive procedure that can be performed only in specialized centers. Alternatively, the model for end-stage liver disease (MELD) has been used as an easily assessed, non-invasive score for prediction of the outcome of patients with cirrhosis (9) supporting the need for liver transplantation in values above the threshold of 15. However, MELD score was first applied for the estimation of 90-day mortality after transjugular intrahepatic portosystemic shunt (TIPS) implementation and not as a general predictor of cirrhotic patients’ outcome (9). Furthermore, its role in compensated patients with lower MELD scores has not been adequately evaluated (10). Currently, liver stiffness measurement (LSM) by elastographic techniques has been found to correlate with HVPG and the presence of esophageal varices (11-17). Moreover, recent studies have shown that LSM can predict liver decompensation and overall survival in patients with cirrhosis (18,19). However, the good correlation between LSM and HVPG seems to be disrupted or become less strong in case of HVPG >12 mmHg (18). Therefore, most investigators combine LSM with other factors, mainly indicative of splenic structural changes, which are considered to express better the advanced stages of PH (20-23). Among them, spleen stiffness measurement (SSM) has been found to correlate well with HVPG (24-26) and being superior to LSM in ruling out patients at risk for variceal bleeding (27). Nonetheless, the number of studies having investigated the role of SSM on the prediction of liver decompensation and survival in patients with cirrhosis is limited (28,29).

The aim of our study was to evaluate if SSM by two-dimension shear-wave elastography (2D-SWE) could predict better the probability of decompensation and survival in cirrhotic patients, compared to LSM and MELD score.

## Material and Methods

Over a 4-year period (January 2018-December 2021), all cirrhotic patients 18-75 years old who attended our outpatient liver clinics were considered eligible for inclusion in the study regardless of the etiology and severity of liver disease.

The diagnosis of cirrhosis was based on clinical and laboratory findings, imaging studies, or liver histology when available. At baseline, all patients underwent LSM, SSM and blood tests including platelet count, prothrombin time, serum albumin, serum creatinine, international normalized ratio (INR), serum aspartate aminotransferase (AST), alanine aminotransferase (ALT), and bilirubin. Likewise, clinical data such as presence of ascites or hepatic encephalopathy (HE) and their severity were noted. The severity of liver disease was determined by Child-Turcotte-Pugh (CTP) and MELD score. After recruitment, clinical and laboratory evaluation was undertaken every 3-6 months and clinical events were recorded. Death, liver transplantation and liver decompensation in previously compensated cirrhotic patients were the end-points of this study. The protocol was approved by the Ethics Committee of General Hospital of Athens “Laiko”, Greece. A written consent was obtained from each patient with respect to all ethical guidelines issued by the 2000 revision (Edinburgh) of the 1975 Declaration of Helsinki.

### Two-dimension SWE (2D-SWE)

All patients underwent LSM and SSM by 2D-SWE performed in fasting patients by a single experienced operator (more than 500 exams). The Aixplorer ultrasound system (Supersonic Imagine S.A., Aix-en-Provence, France) with an abdominal 3.5 MHz curved array probe was used, as recommended (30). LSMs were carried out on the right lobe of the liver through the intercostal spaces with the patient in the supine position and the right arm maximally abducted. The 2D-SWE box was placed in an area of the parenchyma free of large vessels, avoiding liver capsule. SSMs were performed in the supine position with the left arm in maximum abduction and by placing the probe in the left inter-costal spaces. All examinations were performed by a single experienced user (DSK) who had previously completed >500 LSM and SSM by 2D-SWE. LSMs and SSMs were considered reliable when the following criteria were fulfilled: (1) temporal stability of the selected liver area for at least 3 seconds before measurement; (2) two-dimensional quality confirmed by a homogenous color in the region of interest; (3) a measurement region (Q-box) of at least 10 mm. Liver or spleen stiffness failure was defined as either no signal obtained or failure to obtain a reliable 2D-SWE measurement, i.e, no temporal or spatial stability and/or Q-box <10 mm (31,32). Ten reliable LSM and 10 reliable SSM were obtained from each patient and the mean values were respectively calculated. The standard deviation (SD) was <20% of the mean value of LSM and SSM, respectively.

### Statistical analysis

Statistical analysis was done by using SPSS (SPSS software; SPSS Inc, Chicago, IL, USA). Quantitative variables were compared with student’s t-test or Mann-Whitney test for normally and non-normally distributed variables respectively. Qualitative variables were compared with corrected Chi-squared test or two-sided Fisher’s exact test, as appropriate. The relationship between parameters was estimated by using the Spearman’s correlation coefficient. Kaplan-Meier curves were used for the estimation of the probabilities of transplantation free survival, which were compared between subgroups by the log-rank test. Univariable and multivariable Cox regression analyses were performed in order to identify the factors independently associated with liver decompensation and transplantation free survival and hazard ratios (HR) with their 95% confidence interval (CI) are provided. The area under the receiving operating characteristic (AUROC) curves for SSM predictability, as well as sensitivity, specificity, positive predictive value (PPV) and negative predictive value (NPV) were calculated. The c-statistics of AUROC curves were provided with their 95% confidence intervals. Diagnostic accuracy was considered to be poor in case of a c-statistic <0.65, moderate in case of a c-statistic 0.65-0.75, good in case of a c-statistic 0.76-0.85 and excellent in case of a c-statistic >0.85. The optimal cut-off was selected from the AUROCs curves as the point which provided the maximum sum of sensitivity and specificity. All tests were two sided and p values <0.05 were considered to be significant.

## Results

### Patient baseline characteristics

We initially evaluated 196 patients. Nineteen were excluded due to inability to perform SSM. Finally, 177 patients were included in our study. Their main baseline characteristics are presented in **Table 1**. The majority were males (62.7%), whereas their mean age was 57±12 years. The mean duration of follow-up has been 31±18 months. The main causative liver disease was alcoholic liver disease (29.9%), while chronic viral hepatitis was responsible in 29.9%, non-alcoholic fatty liver disease in 18.1%, cholestatic liver diseases in 12.4% and other causes in 9.6% of patients. Of the 177 patients, 58.2% had decompensated cirrhosis. At baseline, ascites was present in 44.1% of patients, while 26.6% of them had a history of HE and 15.8% had a history of variceal bleeding.

**Table 1.** Baseline characteristics of 177 cirrhotic patients.

|  |  |
| --- | --- |
| Sex (males) | 111 (62.7%) |
| Age (years) | 57±12 |
| Body mass index (Kg/m<sup>2</sup>) | 26.4±5.1 |
| Platelets (x 10<sup>9</sup>/L) | 115.8±61.8 |
| Liver stiffness measurement (kPa) | 29.2±12.6 |
| Spleen stiffness measurement (kPa) | 36.6±8.6 |
| Portal vein diameter (cm) | 1.41 ± 0.80 |
| MELD score | 13.7 ± 5.9 |
| Child-Pugh score | 6.1±1.5 |
| Child-Pugh class, A | 79 (44.6%) |
| B | 82 (46.3%) |
| C | 16 (9%) |
| Compensated cirrhosis | 74 (41.8%) |
| Decompensated cirrhosis | 103 (58.2%) |

Quantitative values expressed as mean ± SD.

### Liver decompensation

Among 74 patients with compensated disease, liver decompensation occurred in 30 (40.5%) cases during follow-up. All events of decompensation were established during the first year of follow-up. Patients who developed liver decompensation had no significant differences regarding the age, BMI, LSM, portal vein diameter and MELD score in comparison to those remained in a compensated status. However, they had significantly lower platelet counts (98±43 vs 128±67 × 10^9^ /L; p=0.045) and higher SSM values (39.8±8.6 vs 33.4±9.1 kPa; p=0.004). In multivariable Cox regression analysis, only SSM, was independently associated with the probability of decompensation (HR: 1.063, 95% CI: 1.009-1.120; p=0.021) (**Table 2**). SSM offered an AUROC of 0.710 (p=0.003) for predicting liver decompensation (**Figure 1A**), with the SSM cut-off point of 37 kPa having sensitivity 74.1%, specificity 72.7% and NPV 81.1%. In patients with baseline LSM >20 kPa, the predictive capability of SSM showed an AUROC of 0.730 (p=0.016) (**Figure 1B**), with sensitivity 85.7%, specificity 55.6% and NPV 81.3% for the cut-off point of 37 kPa.

**Table 2.** Factors associated with the probability of liver decompensation in 74 patients with compensated cirrhotic at baseline.

|  | Factors in patients without<br>vs with development of<br>liver decompensation;<br>p value (t-test) | Univariable Cox<br>regression analysis<br>Hazard ratios (95% CI);<br>p value | Multivariable Cox<br>regression analysis<br>Hazard ratios (95% CI);<br>p value |
| --- | --- | --- | --- |
| Age (years) | 56±12 vs 56±14;<br>p=0.828 | 0.978 (0.949-1.007);<br>0.140 |  |
| Body mass<br>index (Kg/m<sup>2</sup>) | 26.8±5.6 vs 27.3±5.4;<br>p=0.697 | 0.985 (0.907-1.000);<br>0.712 |  |
| Platelets (x<br>10<sup>9</sup>/L) | 128.4±66.7 vs 98.1±43.2;<br>**p=0.045** | 0.991 (0.976-1.005);<br>**0.011** | 0.985 (0.0968-1.003);<br>0.111 |
| Liver stiffness<br>(kPa) | 24.9±12.0 vs 27.7±12.8;<br>p=0.331 | 1.043 (1.010-1.077);<br>**0.011** | 1.000 (0.981-1.071);<br>0.190 |
| Spleen<br>stiffness (kPa) | 33.4±9.1 vs 39.8±8.6;<br>**p=0.004** | 1.069 (1.019-1.122);<br>**0.006** | 1.063 (1.009-1.120);<br>**0.021** |
| Portal vein<br>diameter (cm) | 1.28±0.24 vs 1.30±0.28;<br>p=0.767 | 1.128 (0.178-7.155);<br>0.898 |  |
| MELD score | 11.4±5.1 vs 11.8±4.8;<br>p=0.739 | 1.125 (1.022-1.238);<br>**0.016** | 1.095 (0.979-1.024);<br>0.324 |

Quantitative variables are expressed as mean values ± standard deviation; CI: confidence interval.

**Figure 1A.**
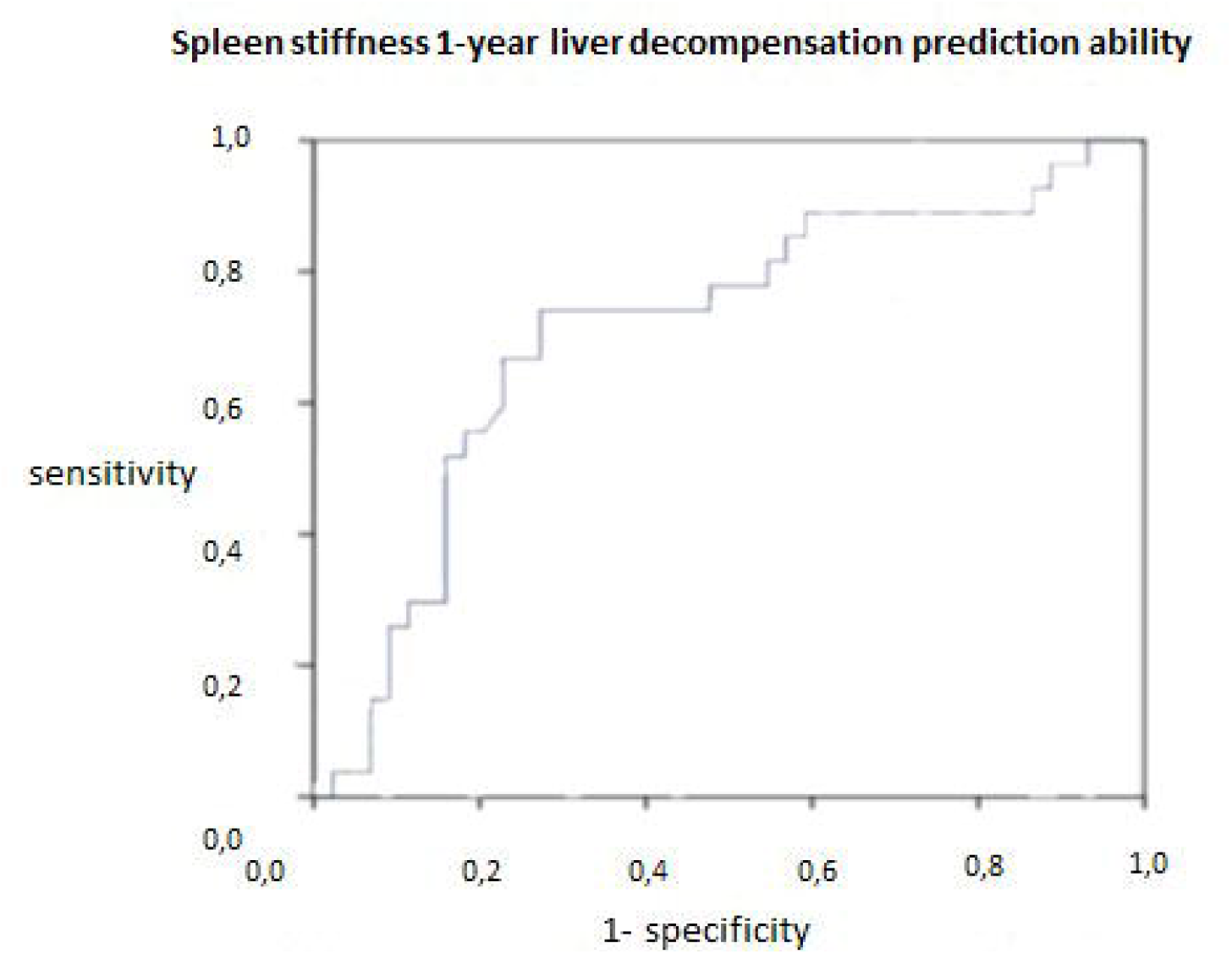
AUROC of spleen stiffness for predicting1-year liver decompensation in 74 patients with baseline compensated cirrhosis.

**Figure 1B.**
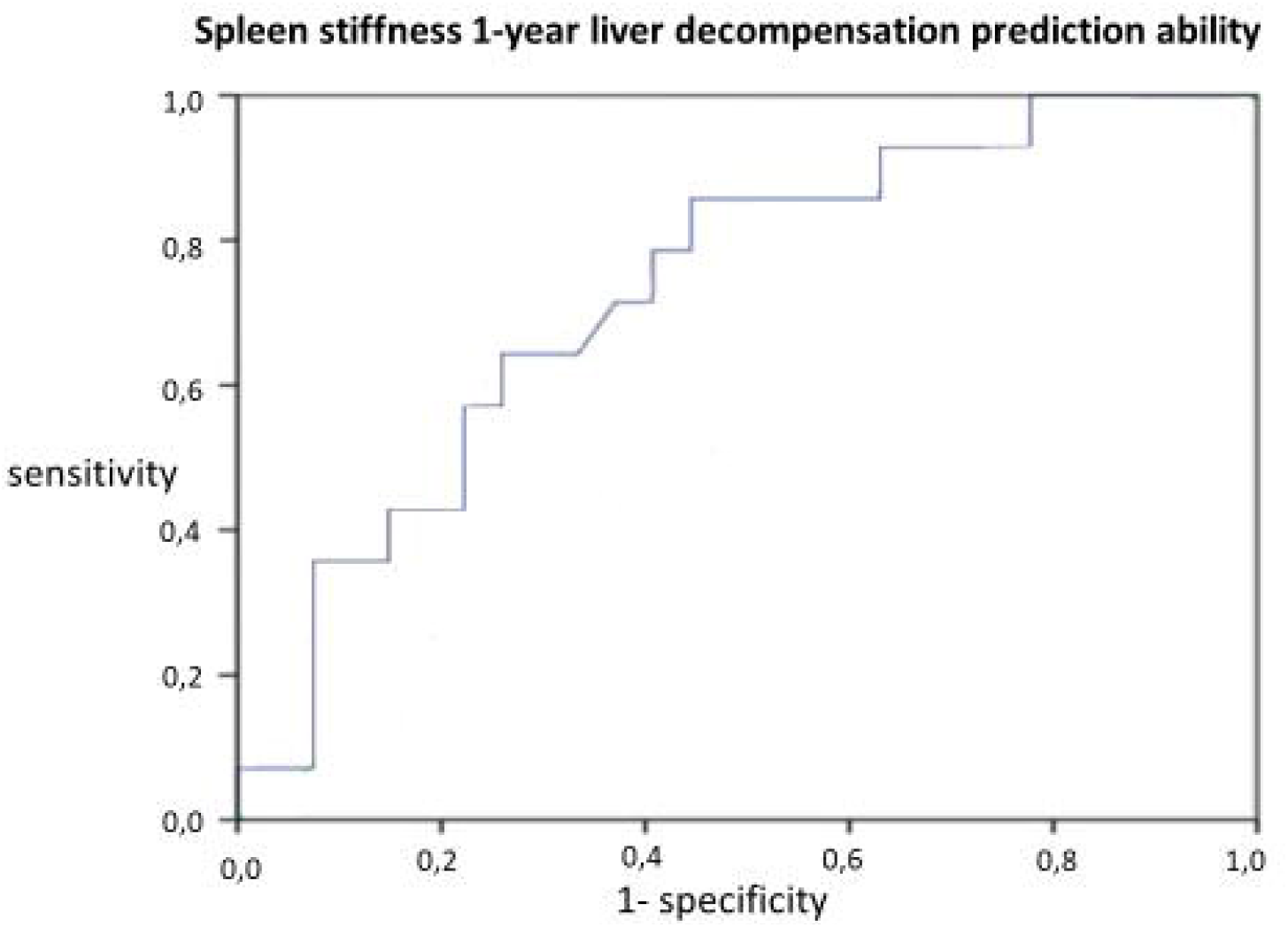
AUROC of spleen stiffness for predicting 1-year liver decompensation in 74 patients with baseline liver stiffness > 20 kPa.

### Survival

During follow-up, 31 (26.6%) of the 177 patients died, whereas another 19 patients (10.7%) underwent liver transplantation. Transplantation free survival was associated with lower baseline SSM (p=0.04), lower MELD (p=0.002) and Child-Pugh score (p=0.010), but not with LSM in univariable analyses **(Table 3)**. In multivariable Cox regression analysis, the occurrence of death or liver transplantation was independently associated only with higher SSM (HR: 1.043; 95% CI:1.003-1.084; p=0.034) (**Table 3**). The AUROC of SSM for predicting 1-year death or liver transplantation was 0.72 (p=0.006) (**Figure 2A**), with the cut-off of 38.8 kPa offering sensitivity 72%, specificity 64% and NPV 95%. The diagnostic performance of SSM to predict the 1-year death or liver transplantation increased in high-risk patients defined by CTP class B or C plus MELD score >10 plus LSM >20 kPa (AUROC: 0.80, p<0.001) (**Figure 2B**). Interestingly, only 1 out of 26 high-risk patients with SSM <38.8 kPa died during the first year of follow-up (NPV: 96.4%).

**Table 3.** Factors associated with the probability of death or liver transplantation in 177 patients with cirrhosis.

|  | Factors in patients who survived vs those who died or were transplanted; p value (t-test) | Univariate Cox regression analysis<br>Hazard ratios (95% CI);<br>p value | Multivariate Cox regression analysis<br>Hazard ratios (95% CI);<br>p value |
| --- | --- | --- | --- |
| Age (years) | 57 ± 12 vs 58 ± 12;<br>p=0.756 | 0.974 (0.944-1.005);<br>0.098 |  |
| Body mass index (Kg/m<sup>2</sup>) | 26.7±5.4 vs 26.3±5.7;<br>p=0.653 | 0.951 (0.898-1.008);<br>0.093 |  |
| Platelets (x 10<sup>9</sup>/L) | 113.4±62.5 vs 117.3±59.4;<br>p=0.745 | 0.998 (0.993-1.003);<br>0.357 |  |
| Liver stiffness (kPa) | 28.3±12.8 vs 31.7±12.7;<br>p=0.164 | 1.015 (0.995-1.036);<br>**0.137** |  |
| Spleen stiffness (kPa) | 35.8±8.6 vs 39.3±8.9;<br>**p=0.004** | 1.052 (1.016-1.089);<br>**0.005** | 1.043 (1.003-1.084);<br>**0.034** |
| Portal vein diameter (cm) | 1.31±0.26 vs 1.38±0.68;<br>p=0.598 | 0.098 (0.764-1.304);<br>0.988 |  |
| MELD score | 12.8±5.8 vs 14.5±5.9;<br>p=0.151 | 1.081 (1.030-1.133);<br>**0.002** | 1.047 (0.981-1.117);<br>0.166 |
| Child-Pugh score | 6.6±1.7 vs 7.1±1.3;<br>p=0.165 | 1.754 (1.143-2.692);<br>**0.010** | 1.078 (0.837-1.386);<br>0.562 |

Quantitative variables are expressed as mean values ± standard deviation; CI: confidence interval.

**Figure 2A.**
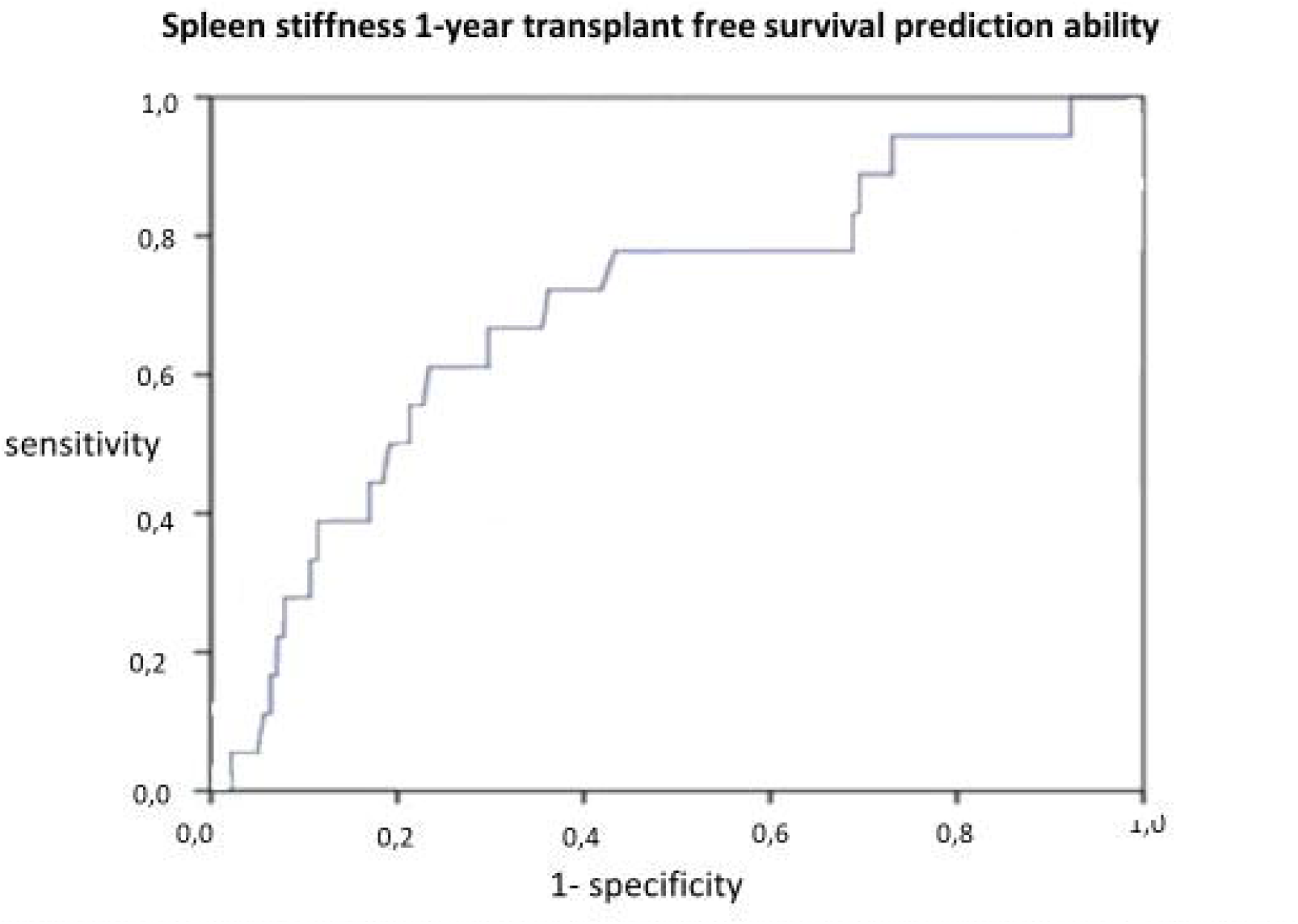
AUROC of spleen stiffness for predicting 1-year transplant free survival in all 177 patients

**Figure 2B.**
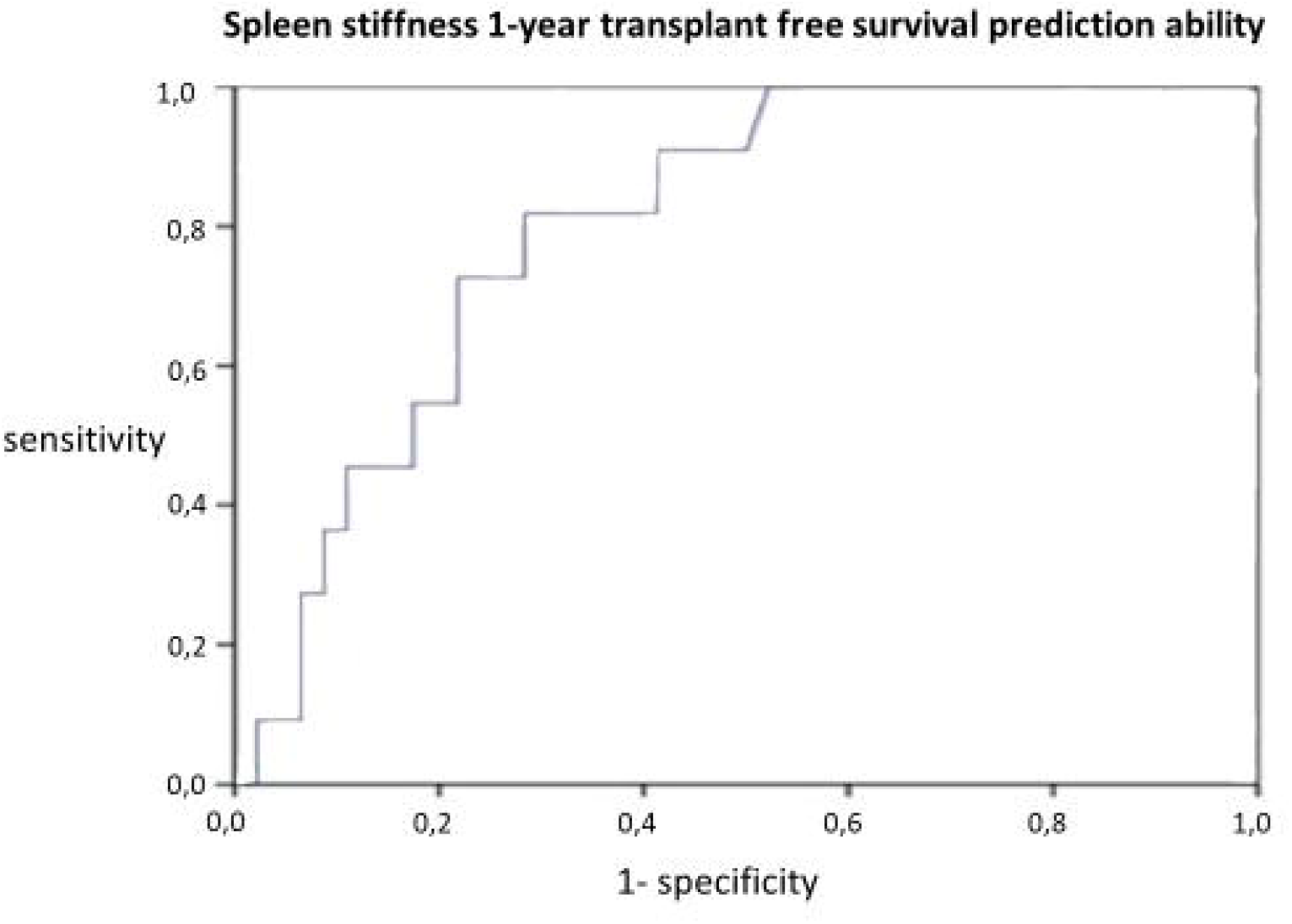
AUROC of spleen stiffness for predicting 1-year transplant free survival in high risk cirrhotic patients defined by Child-Pugh class B or C plus MELD score > 10 plus LSM > 20kPa

In Kaplan-Meier analysis, patients with baseline SSM <38.8 kPa had significantly better mean transplantation free survival compared to those with SSM ≥38.8 kPa (52 vs 42 months, log-rank test: p=0.05) (**Figure 3A**). Similarly, in patients with MELD score >10, cases with baseline SSM <38.8 kPa had significantly better transplantation free survival compared to cases with baseline SSM ≥38.8 kPa (50 vs 39 months, p=0.016) (**Figure 3B**). One the other hand, transplantation free survival did not differ between patients with MELD score ≤10 or >10, patients with LSM ≤20 or >20 kPa, or patients with LSM ≤25 or >25 kPa. Patients with both MELD score >10 plus LSM >25 kPa showed a trend for shorter transplantation free survival compared to patients with both MELD <10 and LSM <25 kPa (45 vs 51 months, p=0.063).

**Figure 3A.**
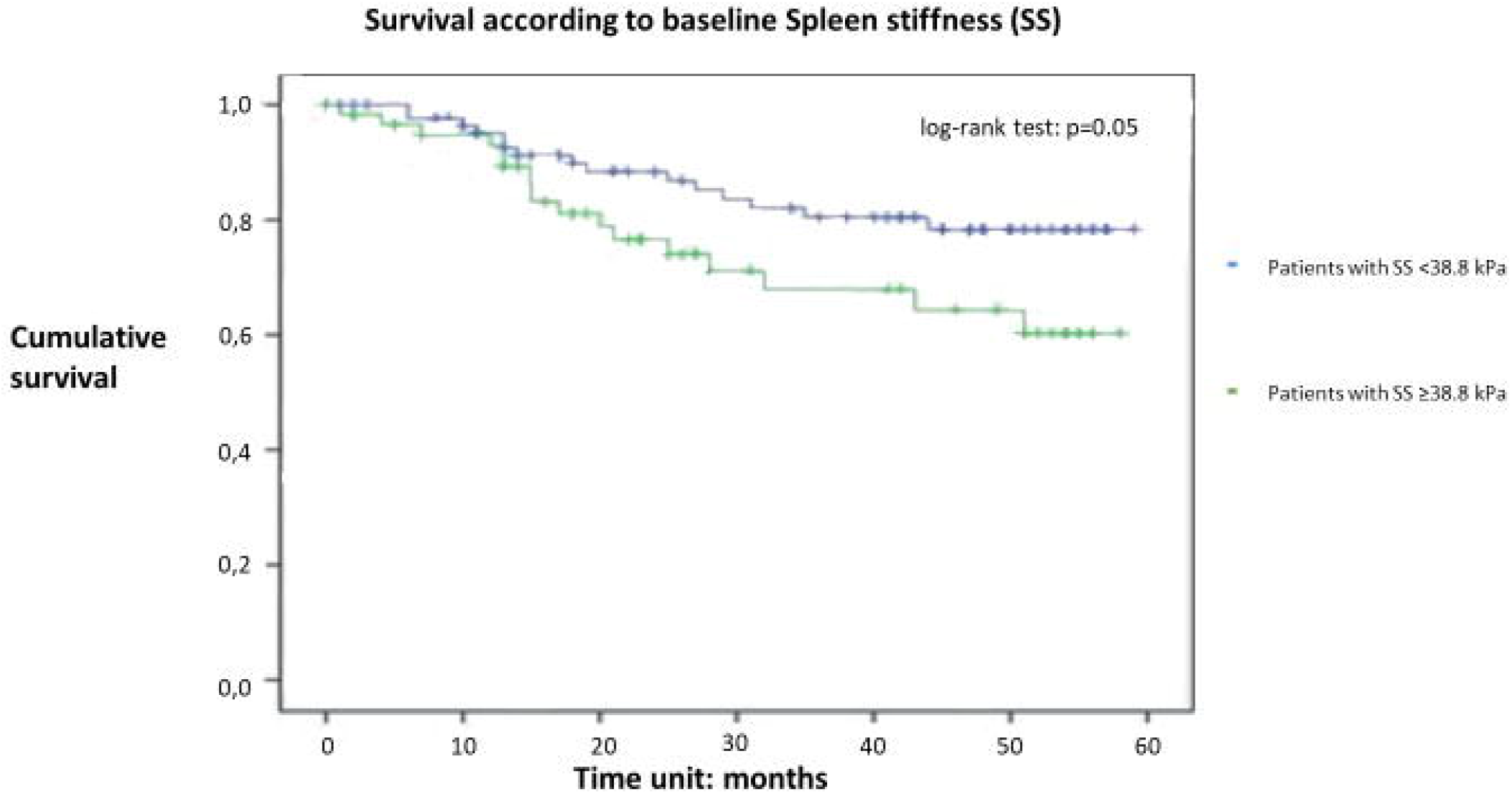
Kaplan-Meier curves of survival rates in all cirrhotic patients in relation to their baseline spleen stiffness (≥38.8 or <38.8kPa)

**Figure 3B.**
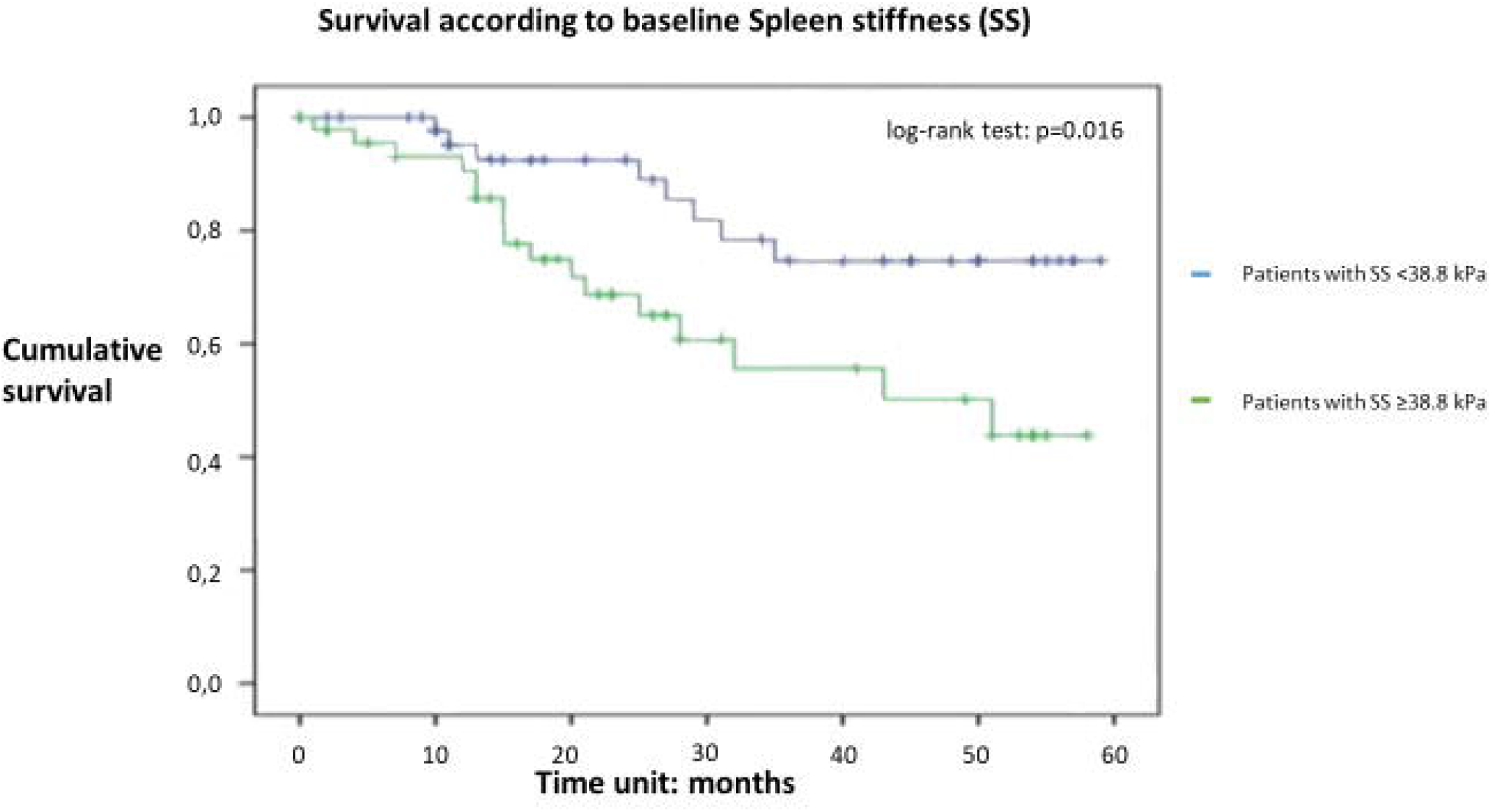
Kaplan-Meier curves of survival rates of patients with baseline MELD score >10 in relation to their baseline spleen stiffness (≥38.8 or <38.8kPa)

## Discussion

Patients with liver cirrhosis represent a large and heterogeneous population with different outcomes associated with the degree of liver failure and the severity of PH at diagnosis. Furthermore, the prognosis may change being affected mostly by the potential of treatment against the cause of liver injury. HVPG, which represents better the severity of PH and predicts well the probability of decompensation and poor outcomes, is an invasive procedure not available in most centers (3,4). On the other hand, MELD score and LSM, which are non-invasive and easy to perform, do not seem to correlate well with the severity of PH. In particular, the significance of MELD score in patients with compensated cirrhosis has not been evaluated, whereas LSM does not seem to align with the advanced stages of PH (10,16,18). Many studies have shown that SSM may express better the more severe phases of PH, compared to LSM. However, most studies investigated the ability of SSM to detect or rule out the presence of high-risk bleeding varices, while data about the accuracy of the method on predicting liver decompensation and/or survival are lacking (24-28). According to our findings, the predictability of SSM is superior compared to MELD score and LSM in patients with baseline compensated cirrhosis since SSM represents the only factor which is independently associated with liver decompensation within the first year of follow-up. SSM seems to have moderate accuracy for predicting liver decompensation in this setting, but the overall 1-year probability of remaining at a compensated stage is approximately 80% in cases with baseline SSM <37 kPa. These results were also confirmed in patients with LSM >20 kPa, who are potentially at increased risk for more severe PH. Indeed, it was found that approximately 80% of such patients will not develop liver decompensation during the next year if their SSM is lower than 37 kPa.

Previous studies have also supported the superiority of SSM as a marker of PH in comparison to LSM, regardless of the etiology of liver cirrhosis (24,33,34). A meta-analysis has shown that esophageal varices, one of the most serious complications of PH, can be detected by SSM with higher sensitivity and specificity compared to LSM (0.88 vs 0.78 and 0.83 vs 0.66, respectively) (35). Furthermore, in studies of patients with compensated cirrhosis of viral etiology, SSM was found to be superior than LSM and similar to HVPG in predicting the first clinical decompensation event (24,28). Recently, Meister et al investigated patients with liver disease of any etiology and reported that those who developed decompensation had SSM >39 kPa (36). In our study, similar results were obtained, as we showed that patients with baseline SSM <37 kPa had <20% probability of developing decompensation during the next year. One difference between our and previous studies is the use of different elastographic methods, as SSM was assessed by transient elastography (TE) in other studies and by 2D-SWE in our study. Although both TE and 2D-SWE are available in our center, we preferred 2D-SWE for this study, as previous reports suggested that 2D-SWE can be used for LSM and SSM in order to rule out the presence of clinically significant PH, showing similar or greater accuracy compared to TE (37,38), while SSM by TE seems considerably difficult to be performed having failure rates higher than LSM (39). For this reason, a novel spleen-dedicated TE probe has been recently developed (40), which however is not available in our center.

Apart from the ability to predict liver decompensation, we showed that SSM was superior to LSM and MELD score for predicting patients’ outcome. In Cox regression analysis, only higher SSM was independently associated with death or liver transplantation. SSM offered a moderate predictability of 1-year death or liver transplanation (AUROC: 0.72), although patients with SSM <38.8 kPa had 95% probability of transplantation-free survival during the next year. Importantly, the predictability of SSM was maintained even among patients potentially being at the highest risk for a poor outcome, i.e. those with Child-Pugh class B or C and MELD >10 and LSM>20 kPa. In particular, 96% of patients with SSM <38.8 kPa survived without liver transplantation during one year of follow-up, while the predictability of SSM increased from moderate to good (AUROC: 0.80) in this subgroup of patients. The findings from Cox regression analysis were in agreement with the results of the Kaplan-Meier curves for survival rates, which were significantly different between patients with baseline SMM <38.8 kPa and ≥38.8 kPa but did not differ in relation to patients’ subgroups according to MELD score or LSM. Specifically, the mean transplantation free survival of patients with baseline SMM <38.8 kPa was approximately 10 months longer than the mean transplantation free survival of patients with baseline SMM ≥38.8 kPa and such a difference was maintained even when only patients with MELD score >10 were considered.

To date, only a small number of studies has reported an association between SSM and survival in cirrhotic patients. However, these studies have investigated the role of SSM as a predictive factor of survival after the insertion of TIPS or postoperatively after hepatic resection for hepatocellular carcinoma (41,42). The association between SSM and mortality in cirrhotic patients not undergoing any invasive intervention was determined prospectively by Takuma et al. In that study, point SWE was used and the cut-off value of 3.43 m/sec identified patients’ death with a 95% NPV and 76% accuracy (29). Our results are in agreement with the above. Moreover, in the present study, we confirmed the predictability of SSM for survival in patients at high risk for major complications, according to the most validated factors (Child-Pugh class, MELD score and LSM), who had 96% probability of transplantation free survival over the next year if their SSM was below 38.8 kPa.

In conclusion, our study demonstrated that SSM by 2D-SWE is superior to LSM and MELD score in predicting liver decompensation and transplantation-free survival of cirrhotic patients. In addition, SSM seemed to be superior even than the combination of MELD score and LSM. If further studies confirm these results, SSM can play a crucial role in order to help physicians to determine a better strategy for preventing liver-related complications and death in patients with cirrhosis.

## Data Availability

All data produced in the present work are contained in the manuscript

## Abbreviations

PH: portal hypertension
HVPG: hepatic venous pressure gradient
MELD: Model for End-stage Liver Disease
TIPS: Transjugular Intrahepatic Portosystemic Shunt
SSM: spleen stiffness measurement
2D-SWE: two-dimension shear-wave elastography
INR: international normalized ratio
AST: aspartate aminotransferase
ALT: alanine aminotransferase
HE: hepatic encephalopathy
CTP: Child-Turcotte-Pugh
SD: standard deviation
AUROC: Area Under the Receiving Operating Characteristic
PPV: positive predictive value
NPV: negative predictive value
HR: hazard ratios
CI: confidence interval
BMI: body mass index
TE: transient elastography.

## Informed consent

Informed consent was obtained from all individual participants included in the study.

## Ethical approval

The study was performed according to the principles of the declaration of Helsinki as revised in 1983 and approval was obtained by the Ethics Committee of General Hospital of Athens “Laiko”, Greece.

## Animal research

Not applicable.

All authors contributed to the interpretation of the data and reviewed and approved the manuscript.

## Figure legends

**Figure 1.** AUROC of spleen stiffness for predicting 1-year liver decompensation in 74 patients with baseline compensated cirrhosis (A), or in those with baseline liver stiffness >20 kPa (B).

**Figure 2.** AUROC of spleen stiffness for predicting 1-year transplant free survival in all 177 patients (A), or in high-risk cirrhotic patients defined by Child-Pugh class B or C plus MELD score >10 plus LSM >20 kPa (B).

**Figure 3.** Kaplan-Meier curves of survival rates in all cirrhotic patients in relation to their baseline spleen stiffness (≥38.8 or <38.8 kPa) (A), or in those with baseline MELD score >10 in relation to their baseline spleen stiffness (≥38.8 or <38.8 kPa) (B).

